# TaME-seq2: Tagmentation-assisted multiplex PCR enrichment sequencing for viral genomic profiling

**DOI:** 10.1101/2022.12.22.22283851

**Authors:** Alexander Hesselberg Løvestad, Milan S. Stosic, Jean-Marc Costanzi, Irene Kraus Christiansen, Hege Vangstein Aamot, Ole Herman Ambur, Trine B. Rounge

**Affiliations:** Faculty of Health Sciences, OsloMet - Oslo Metropolitan University, Oslo, Norway; Department of Microbiology and Infection Control, Akershus University Hospital, Lørenskog, Norway; Department of Clinical Molecular Biology (EpiGen), Division of Medicine, Akershus University Hospital and University of Oslo, Lørenskog, Norway; Department of Research, Cancer Registry of Norway, Oslo, Norway; Centre for Bioinformatics, Department of Pharmacy, University of Oslo, Oslo, Norway

**Keywords:** Intra-host variation, Library preparation, NGS, HPV, SARS-CoV-2, Virology, Genomics, Viral integration

## Abstract

**Background:** Previously developed TaME-seq method for deep sequencing of HPV, allowed simultaneous identification of the HPV DNA consensus sequence, low-frequency variable sites, and chromosomal integration events. The method has been successfully validated and applied to the study of five carcinogenic high-risk (HR) HPV types (HPV16, 18, 31, 33, and 45). Here, we present TaME-seq2 with an updated laboratory workflow and bioinformatics pipeline. The HR-HPV type repertoire was expanded with HPV51, 52, and 59. As a proof-of-concept, TaME-seq2 was applied on SARS-CoV-2 positive samples showing the method’s flexibility to a broader range of viruses, both DNA and RNA.

**Results:** Compared to TaME-seq version 1, the bioinformatics pipeline of TaME-seq2 is approximately 40x faster. In total, 23 HPV-positive samples and seven SARS-CoV-2 clinical samples passed the threshold of 300x mean depth and were submitted to further analysis. The mean number of variable sites per 1000 bp was ∼ 1.5x higher in SARS-CoV-2 than in HPV-positive samples. Reproducibility and repeatability of the method were tested on a subset of samples. A viral integration breakpoint followed by a partial genomic deletion was found in within-run replicates of HPV59-positive sample. Identified viral consensus sequence in two separate runs was >99.9 % identical between replicates, differing by a couple of nucleotides identified in only one of the replicates. Conversely, the number of identical minor nucleotide variants (MNVs) differed greatly between replicates, probably caused by PCR-introduced bias. The total number of detected MNVs, calculated gene variability and mutational signature analysis, were unaffected by the sequencing run.

**Conclusion:** TaME-seq2 proved well suited for consensus sequence identification, and the detection of low-frequency viral genome variation and viral-chromosomal integrations. The repertoire of TaME-seq2 now encompasses seven HR-HPV types. Our goal is to further include all HR-HPV types in the TaME-seq2 repertoire. Moreover, with a minor modification of previously developed primers, the same method was successfully applied for the analysis of SARS-CoV-2 positive samples, implying the ease of adapting TaME-seq2 to other viruses.

## 1. Background

New sequencing methods have enabled deep diving into viral genomics and viral-host interactions. Nearly all cases of cervical cancer have persistent infections with human papillomavirus (HPV) as a causative agent^1^. Recent studies of HPV intra-host variation have revealed the presence of minor nucleotide variants (MNVs) below the consensus sequence level that can be of clinical relevance for the development of HPV-induced cervical cancer^2,3^. Additionally, the clinical relevance of intra-host MNVs is not limited to HPV infections, as shown in previous deep-sequencing studies of other viral infections^4–7^. Furthermore, the integration of the HPV genome into the human genome is frequently observed^8^ and is considered a driving event in HPV-induced cancer development^9^.

A tagmentation-assisted multiplex PCR enrichment sequencing protocol, TaME-seq, for deep sequencing of HPV, has previously been developed ^10^. This protocol allows simultaneous identification of the consensus sequence, low-frequency variable sites, and chromosomal integration events within clinical samples. Similar methods generally allow the analysis of either HPV genomic variability or integrations and are also less cost-efficient^11–13^. TaME-seq has been successfully applied to study high-risk (HR) HPV types, HPV16, 18, 31, 33, and 45^10,14,15^. This study presents TaME-seq2 with an updated laboratory workflow and bioinformatics pipeline (Figure 1). In addition, the HPV repertoire is expanded to include three HR-HPV types, HPV51, 52, and 59. Furthermore, we present a proof-of-concept that TaME-seq2 can be used for SARS-CoV-2 sequencing and easily be applied to both DNA and RNA viruses.

**Figure 1.**
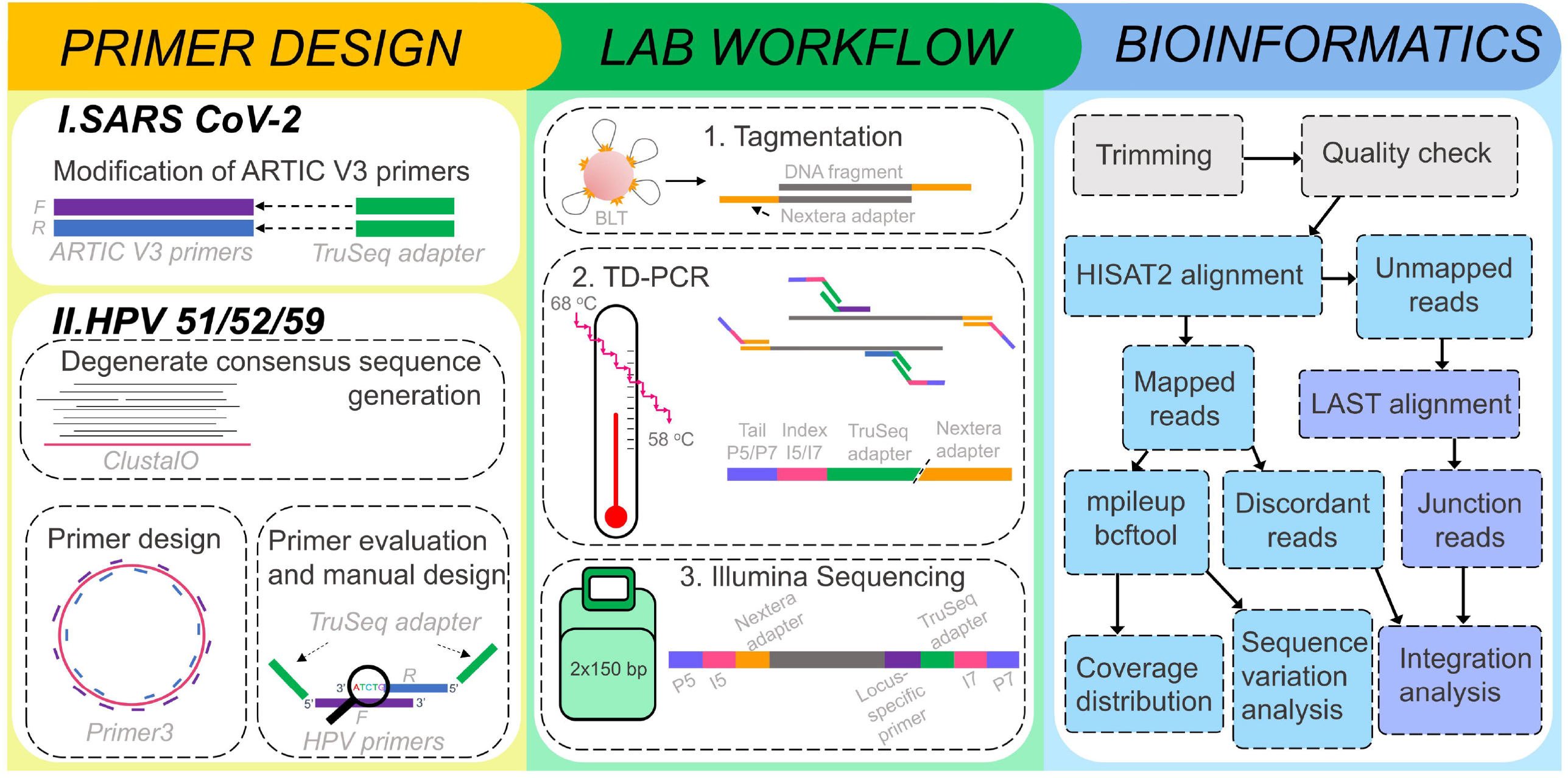
Overview of the TaME-seq2 workflow. The workflow is divided into three steps, primer design shown separately for SARS-CoV-2 and HPV51/52/59, lab workflow consisting of tagmentation step, touch-down PCR and Illumina sequencing, and bioinformatics analysis represented as a flowchart.

## 2. Results

Summarized sequencing results for each virus type are shown in Table 1. Detailed sequencing results of 36 cervical cell samples positive for HPV51/52/59 (12 per type), HPV harbouring plasmid samples with targeted types (two per type), 27 SARS-CoV-2 positive samples, and negative controls are presented in Table S2. The total number of generated raw reads from all HPV samples was 553.3 million, of which 72.9 million mapped to HPV. The percentage of reads mapping to the target HPV was 29.9 % for all samples, but after the low-quality samples were filtered out (three HPV51, five HPV52, and five HPV59), the percentage increased to 40.5 %. Only 0.4 % of the HPV reads mapped to off-target HPV types. No significant difference in the number of off-target mapping reads was found between samples with single and multiple HPV infections (Wilcoxson test, p=0.15). Mean sequencing depth ranged between 0.06x and 82 704x. In total, 23 HPV-positive samples, excluding positive plasmid controls, passed the threshold of 300x mean depth and were submitted to further analysis.

**Table 1.**
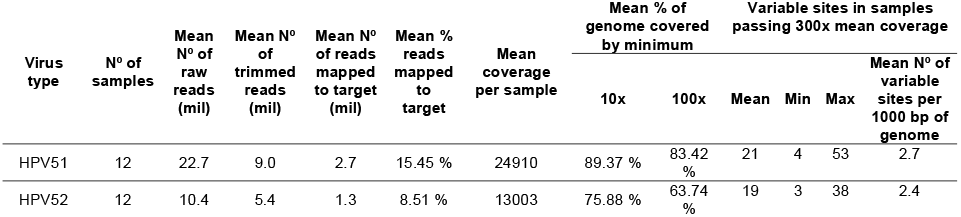
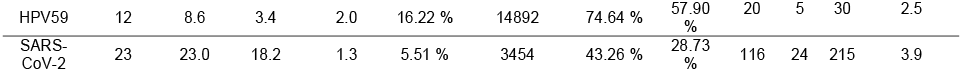
Overview of the sequencing results of HPV 51/52/59 positive cervical-cell samples and SARS-CoV-2 positive samples. A summary of variable sites in clinical samples passing the threshold of 300x mean coverage is also presented.

| Virus type | Nº of samples | Mean Nº of raw reads (mil) | Mean Nº of trimmed reads (mil) | Mean Nº of reads mapped to target (mil) | Mean % reads mapped to target | Mean coverage per sample | Mean % of genome covered by minimum |  | Variable sites in samples passing 300x mean coverage |  |  |  |
| --- | --- | --- | --- | --- | --- | --- | --- | --- | --- | --- | --- | --- |
|  |  |  |  |  |  |  | 10x | 100x | Mean | Min | Max | Mean Nº of variable sites per 1000 bp of genome |
| HPV51 | 12 | 22.7 | 9.0 | 2.7 | 15.45 % | 24910 | 89.37 % | 83.42 % | 21 | 4 | 53 | 2.7 |
| HPV52 | 12 | 10.4 | 5.4 | 1.3 | 8.51 % | 13003 | 75.88 % | 63.74 % | 19 | 3 | 38 | 2.4 |
| HPV59 | 12 | 8.6 | 3.4 | 2.0 | 16.22 % | 14892 | 74.64 % | 57.90 % | 20 | 5 | 30 | 2.5 |
| SARS-CoV-2 | 23 | 23.0 | 18.2 | 1.3 | 5.51 % | 3454 | 43.26 % | 28.73 % | 116 | 24 | 215 | 3.9 |

Sequencing of SARS-CoV-2 positive samples resulted in 529.5 million raw-reads, of which 28.8 million mapped to the SARS-CoV-2 genome. Seven samples passed the minimum requirement for subsequent analysis, with a percentage of reads mapping to SARS-CoV-2 being 56.1 (Table S2). Coverage plots of three representative samples from each HPV type and one SARS-CoV-2 positive sample are shown in Figure 2.

**Figure 2.**
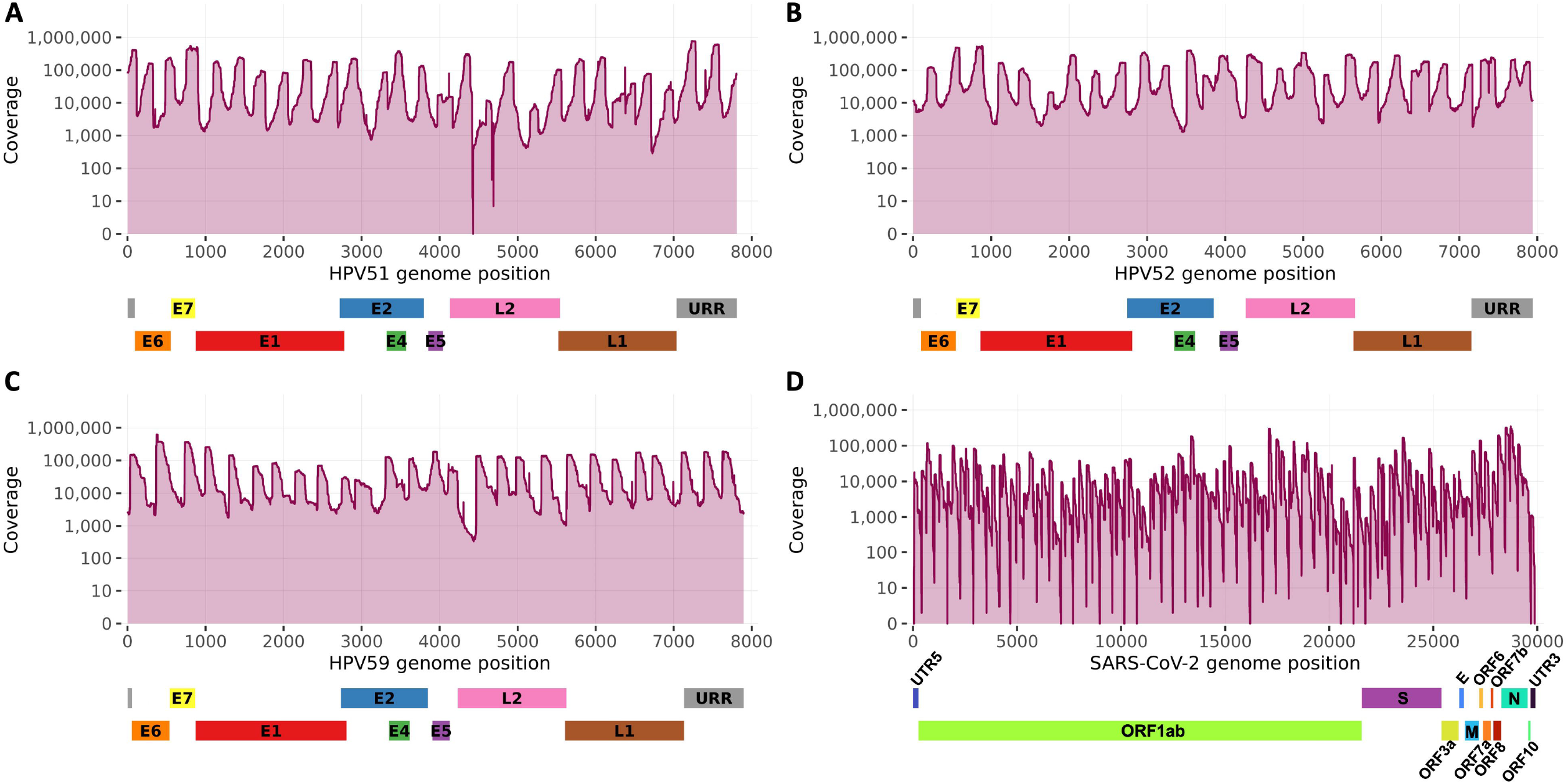
Sequencing coverage in representative HPV51, 52, 59, and SARS-CoV-2 positive samples. The coverage plots of A) HPV51, B) HPV52, C) HPV59, and D) SARS-CoV-2 aligned to the respective target HPV and SARS-CoV-2 genomes.

### 2.1. Analysis of viral deletion and viral integration into the human genome

Integration was found in only one within-run replicate 10a-HPV59/10b-HPV59. Two reported integration breakpoints were found in the HPV genes E2 and E5, and in the human chromosome 2 (Table S3, Figure S2). Deletions were found between 2348 bp – 3039 bp in 10a-HPV59, and between 2349 bp – 3036 bp in 10b-HPV59 corresponding to the segments of E1 and E2 genes (Figure S1). In SARS-CoV-2 positive samples, neither viral deletion nor integration events were detected.

Moreover, two short regions with a coverage drop were observed in all HPV51 samples. The affected nucleotides positions were found between 4416-4430 and 4690-4692, corresponding to the L2 gene region, most likely due to suboptimal primer hybridization and/or poor alignment against the reference genome (Figure 2A).

### 2.2. Minor nucleotide variation and mutational signature analysis

Minor nucleotide variation analysis identified 21, 19, and 20 average MNVs per sample in HPV51, 52, and 59 positive samples, respectively. The number of MNVs found within individual samples ranged from 3 to 53 irrespective of HPV type (Table 1, Figure S3A). Moreover, MNVs’ positions were scattered throughout the genomes, with HPV59 E7 showing the highest degree of variation per base pair (Figure 3A).

**Figure 3.**
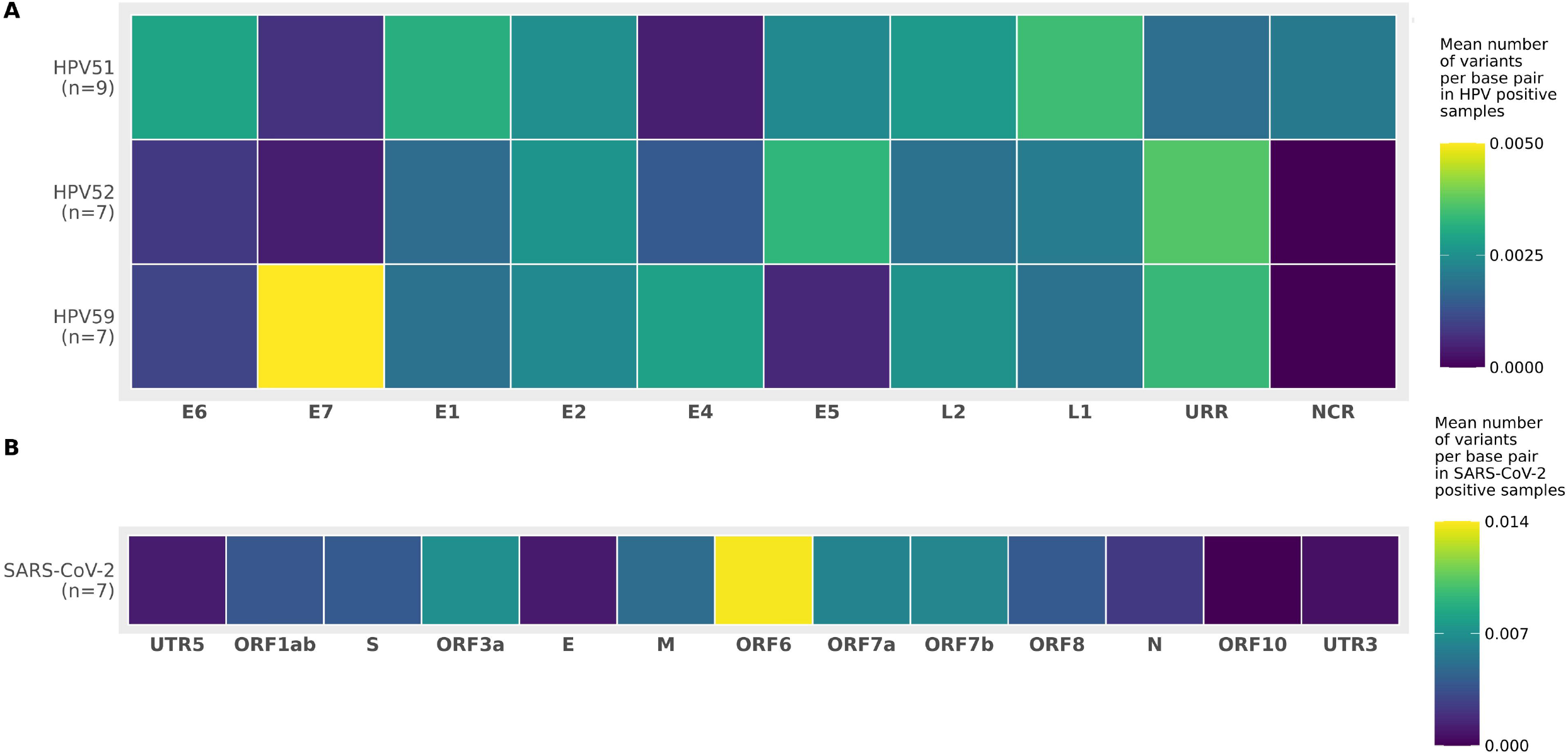
Mean number of variants per gene normalized by gene length presented as heatmap. A) HPV51, HPV52, and HPV59 positives samples B) SARS-CoV-2 positive samples.

The mean number of variable sites in SARS-CoV-2 was 116 (Table 1, Figure S3B). Accounting for gene length, ORF6 had the highest number of MNVs per base pair (Figure 3B). The mean number of variable sites per 1000 bp in SARS-CoV-2 was ∼ 1.5x higher compared to HPV types (Table 1).

Mutational signatures found in HPV51, 52, and 59 were almost identical, with C>T and T>C being the most prevalent substitutions regardless of the trinucleotide context and HPV type (Figure 4). Out of all C>T substitutions detected, the ones in the trinucleotide context TCW (W is either A or T) were the predominant C>T substitutions in HPV51 samples. TCW trinucleotide context has previously been described as a target sequence of APOBEC3 proteins^24^. T>C and C>A substitutions were the most prevalent in SARS-CoV-2 samples (Figure 4).

**Figure 4.**
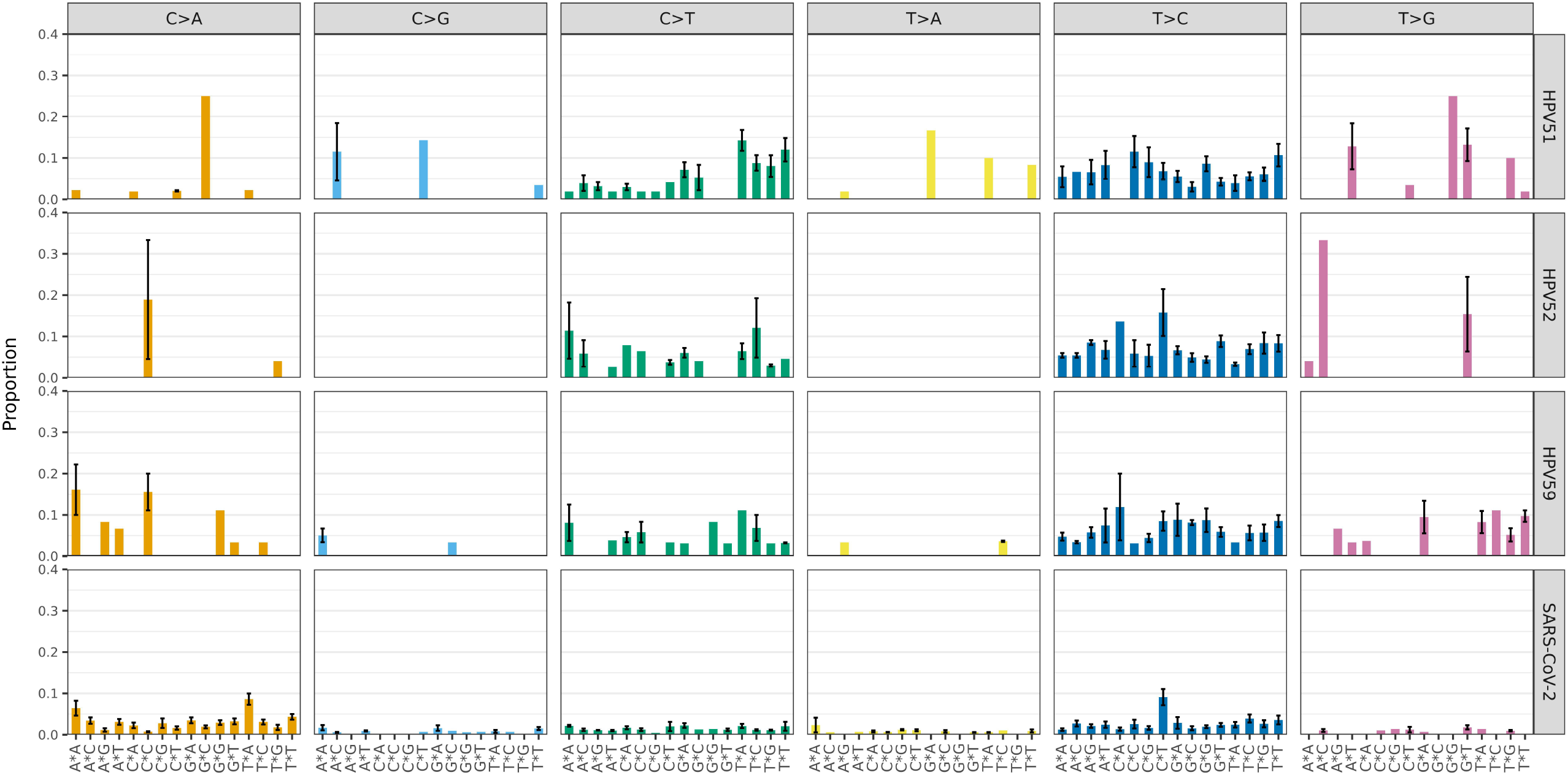
Mutational signatures in HPV51, HPV52, HPV59, and SARS-CoV-2 positive clinical samples. The mean proportion of 96 trinucleotide substitution types is shown below the plots across the different diagnostic categories. Error bars represent the standard error of the mean.

### 2.3. Reproducibility of consensus sequences, MNVs, and mutational signatures between replicates

Technical between-run replicates of six HPV51 and nine HPV52 positive samples were used to assess the reproducibility of both consensus sequence identification and minor nucleotide calling. Replicates were tagmented, PCR amplified, and sequenced independently. In total, seven replicates did not meet the minimum requirements to be included in the analysis due to mean coverage being <300x in one or both samples (n=5) or having failed forward or reverse reactions in one of the samples (n=2).

The sequencing results of the eight replicates included in the analysis are presented in Table 2. The results of two sequencing runs were significantly different (Wilcoxon paired two-tail test) in terms of the number of trimmed reads (p=0.008), reads mapping to HPV (p=0.008), reads mapping to the targeted HPV (p=0.008), and mean coverage between pairs of samples (p=0.008). However, the percentage of genome covered with >100x and >300x was unaffected by the sequencing run (Wilcoxon paired two-tail test, p=0.2, and p=0.5, respectively).

**Table 2.**
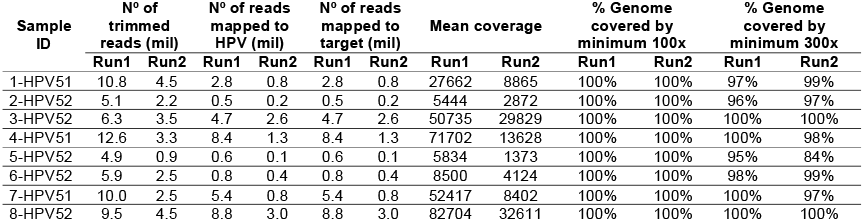
Overview of the results of two sequencing runs of eight HPV-positive replicated samples.

| Sample ID | N° of trimmed reads (mil) |  | N° of reads mapped to HPV (mil) |  | N° of reads mapped to target (mil) |  | Mean coverage |  | % Genome covered by minimum 100x |  | % Genome covered by minimum 300x |  |
| --- | --- | --- | --- | --- | --- | --- | --- | --- | --- | --- | --- | --- |
|  | Run1 | Run2 | Run1 | Run2 | Run1 | Run2 | Run1 | Run2 | Run1 | Run2 | Run1 | Run2 |
| 1-HPV51 | 10.8 | 4.5 | 2.8 | 0.8 | 2.8 | 0.8 | 27662 | 8865 | 100% | 100% | 97% | 99% |
| 2-HPV52 | 5.1 | 2.2 | 0.5 | 0.2 | 0.5 | 0.2 | 5444 | 2872 | 100% | 100% | 96% | 97% |
| 3-HPV52 | 6.3 | 3.5 | 4.7 | 2.6 | 4.7 | 2.6 | 50735 | 29829 | 100% | 100% | 100% | 100% |
| 4-HPV51 | 12.6 | 3.3 | 8.4 | 1.3 | 8.4 | 1.3 | 71702 | 13628 | 100% | 100% | 100% | 98% |
| 5-HPV52 | 4.9 | 0.9 | 0.6 | 0.1 | 0.6 | 0.1 | 5834 | 1373 | 100% | 100% | 95% | 84% |
| 6-HPV52 | 5.9 | 2.5 | 0.8 | 0.4 | 0.8 | 0.4 | 8500 | 4124 | 100% | 100% | 98% | 99% |
| 7-HPV51 | 10.0 | 2.5 | 5.4 | 0.8 | 5.4 | 0.8 | 52417 | 8402 | 100% | 100% | 100% | 97% |
| 8-HPV52 | 9.5 | 4.5 | 8.8 | 3.0 | 8.8 | 3.0 | 82704 | 32611 | 100% | 100% | 100% | 100% |

Consensus sequences between replicates were identical, except for a few positions, ranging between 1-6 nucleotides (Table 3). Conversely, replicates exhibited a large difference in MNVs between the two runs (Table 4). Identical MNVs shared between replicates ranged between zero and three. However, the total number of MNVs in replicates detected between the two runs was not significantly different (Wilcoxon paired two-tail test, p=0.3).

**Table 3.**
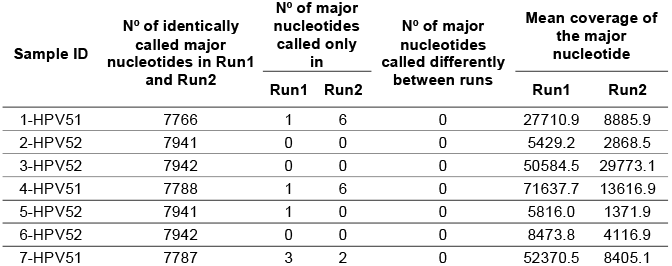
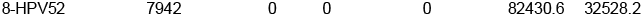
Overview of major nucleotide calling results used to generate consensus genome sequences in eight HPV replicates. For each pair of samples, the number of identical major nucleotides between runs, the number of nucleotides called in one of the runs, and the number of called nucleotides differing between runs are shown. The mean coverage of the called major nucleotides in two runs is also presented.

**Table 4.**
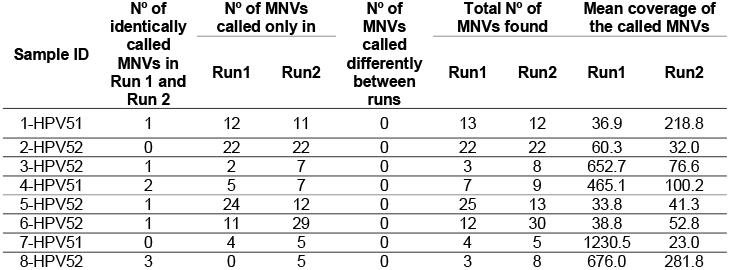
Overview of MNV calling results in eight HPV replicates. For each pair of samples, the number of identical MNVs between runs, the number of MNVs called only in one of the runs, the number of MNVs differing between runs is shown, and the total number of MNVs found. Mean coverage of the called MNVs in two runs is also presented.

| Sample ID | N° of identically called MNVs in Run 1 and Run 2 | N° of MNVs called only in |  | N° of MNVs called differently between runs | Total N° of MNVs found |  | Mean coverage of the called MNVs |  |
| --- | --- | --- | --- | --- | --- | --- | --- | --- |
|  |  | Run1 | Run2 |  | Run1 | Run2 | Run1 | Run2 |
| 1-HPV51 | 1 | 12 | 11 | 0 | 13 | 12 | 36.9 | 218.8 |
| 2-HPV52 | 0 | 22 | 22 | 0 | 22 | 22 | 60.3 | 32.0 |
| 3-HPV52 | 1 | 2 | 7 | 0 | 3 | 8 | 652.7 | 76.6 |
| 4-HPV51 | 2 | 5 | 7 | 0 | 7 | 9 | 465.1 | 100.2 |
| 5-HPV52 | 1 | 24 | 12 | 0 | 25 | 13 | 33.8 | 41.3 |
| 6-HPV52 | 1 | 11 | 29 | 0 | 12 | 30 | 38.8 | 52.8 |
| 7-HPV51 | 0 | 4 | 5 | 0 | 4 | 5 | 1230.5 | 23.0 |
| 8-HPV52 | 3 | 0 | 5 | 0 | 3 | 8 | 676.0 | 281.8 |

To further investigate whether the observed difference would affect the calculation of gene variability, MNVs in different runs were grouped by the genes they occurred in, regardless of the HPV type, and counted (Table S4). Even though MNVs were at different positions, their numbers per gene were not significantly different between runs (Wilcoxon paired two-tailed test, p=0.4).

Furthermore, the reproducibility of mutational signature analysis was also assessed. In both sequencing runs, regardless of the HPV type, the same pattern of C>T and T>C substitutions was detected (Figure S4). The number of all observed substitutions in their respective trinucleotide context was not significantly different between runs, as well as their mean proportions (Wilcoxon paired two-tailed test, p=0.4).

## 3. Discussion

TaME-seq2 enables in-depth analysis of DNA and RNA viruses by exploiting the latest kits and efficient software. Compared to the previous version of TaME-seq, the bioinformatics pipeline is approximately 40x faster without affecting the analysis quality. The laboratory workflow was improved by implementing the latest Nextera tagmentation kit accompanied by touchdown PCR, known to increase the amplification yield and decrease off-target amplification^25^. Adding unique dual indexes minimizes the risk of calling erroneous low-frequency variants due to index hopping.

The number of HPV samples passing the threshold is affected by lower initial DNA input and/or low viral load in samples, a general feature of multiplex target enrichment protocols and reported for the previous version of the protocol^15^. In SARS-CoV-2 samples, the high number of Illumina-tail-extended primers was most likely responsible for the primer-dimer formation, in turn causing reduced amplification yield. Even though the method was successfully applied as a proof-of-concept, further optimization of the laboratory workflow and primer pools is recommended.

The only deletion and viral integration into the human genome were found in the HPV59 positive within-run replicate, indicating a repeatability in detecting deletion/integration events as shown for the previous iteration of the protocol^10^. Detected breakpoints in early genes accompanied by complete or partial deletion of E1 and E2 have frequently been observed for HPV integrations^12^. Integrations into the human genome were not found in SARS-CoV-2 positive samples, confirming previous findings^26^.

The mean number of MNVs in included HPV types was similar and in line with previous studies^14,27–30^. In SARS-CoV-2 samples, a higher mean number of detected MNVs compared to HPV-positive samples is probably the consequence of the significant difference in genome size between HPV and SARS-CoV-2. Furthermore, a higher mutation rate is expected in RNA viruses, which replicate using low-fidelity RNA-dependent RNA polymerase^31^. As the SARS-CoV-2 samples exhibited somewhat lower mean coverage and sequencing depth compared to HPV, it can be expected that higher sequencing coverage would increase the number of detected MNVs.

Mutational signatures found in HPV51, 52, and 59 were almost identical, with C>T and T>C being the most prevalent substitutions regardless of the trinucleotide context and HPV type. The same signatures were also found in previously assessed types HPV16, 18, 31, 33, and 45^14,27^. T>C and C>A substitutions were the most prevalent in SARS-CoV-2 samples, as shown in similar studies^6,7^.

The reproducibility of the TaME-seq2 performance was assessed with eight HPV-positive replicates, which underwent independent library preparation and sequencing. As the number of samples on the flow cells differed between the sequencing runs, the number of generated reads between runs differed. However, the overall performance of TaME-seq2 in terms of the percentage of genome covered with >300x per sample replicate was not significantly different between the two runs.

Consensus sequences were identical between the two runs irrespective of HPV types, except for a few low coverage positions in some of the samples. On the other hand, the shared number of identical MNVs between replicates of each pair was low. Identification of low-frequency variants depends mainly on the coverage at the given site. In our previous study, the number of detected low-frequency MNVs increased with the mean coverage, reaching a saturation plateau at ∼12 000x^10^. As identical nucleotides in replicated samples might have different coverage, the number of detected MNVs at identical positions is expected to differ. Moreover, only a small proportion of HPV genomes in the sample harbours MNVs. As tagmentation, PCR amplification, and sequencing are stochastic processes favouring higher-frequency variants within samples, many low-frequency variants can be expected to stay undetected. Especially PCR amplification introduces biases, favouring amplification of higher-frequency variants^32^, thereby altering the analysed intra-host variation composition in a stochastic manner. Even if the sequencing depth of the whole HPV genome is above the saturation plateau, it cannot be expected that all identical MNVs would be detected after the sample replicates are tagmented, PCR-amplified, and sequenced independently.

The total number of detected MNVs between replicates sequenced in two different runs was not significantly different. Regardless of HPV type, the gene variability did not differ significantly between independent runs. Furthermore, assigning the MNVs by their substitution type in a trinucleotide context did not show a significant difference between runs or HPV types (Figure S4). Overall, the results indicate that the method is suitable for investigating intra-host MNV diversity and is in line with similar studies^2,3^. The mutational signature analysis is robustly detecting the specific within-patient mutation patterns.

## 4. Conclusion

TaME-seq2 proved well suited for both consensus sequence identification, low-frequency viral genome variation, and viral-chromosomal integration analysis. Now, the repertoire of TaME-seq2 encompasses HPV51, 52, and 59 in addition to HPV16, 18, 31, 33, and 45. Our goal is to include all HR-HPV types in the TaME-seq2 repertoire. Moreover, with the addition of Illumina TruSeq-compatible adapter tails to previously developed primers, the same method was successfully applied as a proof-of-concept for the analysis of SARS-CoV-2, implying the ease of adapting TaME-seq2 to a broader variety of viruses. However, further optimization of the laboratory workflow and primer pools is recommended.

## 5. Methods

### 5.1. Sample material

The study included the following material: i) Cervical cell samples positive for HPV51, 52, and 59 (Table S1). The samples are part of a research biobank and were collected between 2005-2008 from women participating in the cervical cancer screening program in Norway^16,17^. These samples had either a single infection or multiple infections with at least one of the targeted types (n=36, 12 per type). ii) Paired technical HPV-positive replicates (n=16), of which 15 pairs were sequenced in separate sequencing runs and one replicate pair sequenced within the same sequencing run. iii) Positive controls, two HPV harbouring plasmid samples per targeted type from the Equalis global HPV DNA typing proficiency study 2019^18^ containing either a single HPV type or multiple HPV types. iv) 27 SARS-CoV-2 cDNA samples from a study on suspected intra-hospital SARS-CoV-2 transmissions during the first wave of the COVID-19 pandemic^19^.

### 5.2. Laboratory workflow

The updated TaME-seq2 protocol includes a previously described protocol used for designing HPV51, 52, and 59 specific primers^10^ (Figure 1). SARS-CoV-2 primers were designed by adding Illumina TruSeq-compatible adapter tails to the 5’ends of the ARTIC Version 3 primer set designed by the Artic Network^20^.

TaME-seq2 compatible unique dual indexes were designed using the IDT® for Illumina® DNA/RNA UD Indexes as a template and adding Nextera-compatible adapter tails as described previously^10,21^. All primers were synthesised by Thermo Fisher Scientific, Inc. (Waltham, MA).

HPV and SARS-CoV-2 specific forward and reverse primer pools were prepared separately in equal volumes and diluted to a concentration of 15 μM. The unique dual indexes were diluted to 10 μM and corresponding i5/i7 pairs were combined.

The updated TaME-seq2 protocol includes tagmentation and post-tagmentation clean-up using the Illumina® DNA Prep Tagmentation kit (Illumina, Inc., San Diego, CA) according to the manufacturer’s recommendations but using half the recommended reaction volumes. Qiagen Multiplex PCR Master Mix (Qiagen, Hilden, Germany) was prepared and mixed with tagmented DNA still attached to the bead-linked transposomes. DNA-containing master mixes were divided into two separate PCR reactions of equal volumes, and forward or reverse virus-specific primer pools were added to the respective PCR reactions. Primer pool concentrations were 0.6 μM and the final concentration of i5/i7 index primers was 0.8 μM per 25 μl PCR reaction.

The updated protocol includes a touchdown PCR consisting of 5 minutes initial denaturation and hot start at 95 °C; 10 touchdown cycles consisting of denaturation at 95 °C for 30 seconds, annealing at 68 °C for 90 seconds, decreasing by 1 °C per cycle, and elongation at 72 °C for 30 seconds; 26 cycles with the fixed annealing temperature at 58 °C for 90 seconds with the PCR cycling parameters as in the previous cycling step; and final extension at 68 °C for 10 minutes. The bead-linked transposomes were removed before the forward and reverse reactions were pooled in equal volumes and submitted to clean-up and two-sided size selection using purification beads (Illumina® DNA Prep Tagmentation kit) according to the manufacturer’s instructions. The libraries were then washed three times using a 0,65x ratio of Sample Purification beads (Beckman Coulter, Brea, CA) to remove excess primer-dimers and short fragments <300 bp.

Due to the detection of primer-dimer excess (⍰150bp) in the SARS-CoV-2 sample libraries after size selection and clean-up, a DNA gel-extraction was performed using the Wizard® SV Gel and PCR Clean-Up System (Promega, Madison, WI) to extract fragments >300 bps.

Quality and quantity of pooled libraries were assessed on Agilent 2100 Bioanalyzer using Agilent High Sensitivity DNA Kit (Agilent Technologies Inc., Santa Clara, CA) before sequencing on the NovaSeq 6000 platform with the SP Reagent Kit v1.5 (Illumina, Inc., San Diego, CA). Samples were sequenced as 151 bp paired-end reads.

### 5.3. Bioinformatic workflow

The TaME-seq2 bioinformatic pipeline includes trimming of raw pair-end reads by removal of adapters, virus-specific primers, and nucleotides with a quality < 20 by cutadapt (v3.4) and quality check by FastQC (v.0.11.9) and MultiQC (v1.10.1). The trimmed reads were mapped to the virus-specific and human (hg38) reference genomes using HISAT2^22^ (v2.2.1). All available HPV reference genomes retrieved from the Papillomavirus Episteme (PaVE)^23^ database were used in the reference genome file for HPV-positive samples, while NC_045512.2 was used for SARS-CoV-2 positive samples. Subsequently, mpileup from bcftool (v1.12) compiled the mapping statistics at a single nucleotide resolution. Mapping statistics and sequencing coverage from forward and reverse reactions for each sample were combined and visualised with an in-house R (v3.5.1) script, enabling evaluation of the method performance and downstream chromosomal integration and sequence variation analysis. This study excluded samples with <300x mean sequencing depth from downstream analyses. This threshold may vary depending on the research aim.

### 5.4. Integration analysis

Viral chromosomal integration analysis was performed as described previously^10^. In brief, discordant read-pairs from the HISAT2 mapping were identified, and potential integration sites were reported if ≥2 human-mapping reads exhibited unique start and/or end coordinates. The LAST aligner remapped HISAT2-unmapped reads to the reference genomes, thereby identifying junction reads. Positions covered with ≥3 junction reads, with unique start and end coordinates, were designated potential integration breakpoints. All viral-human integration points were validated by visualizing junction and discordant reads mapped to the identified human genome regions by Integrative Genomics Viewer (IGV, v2.5.3). Integration sites reported exclusively by read mapping to repetitive regions, split reads, and reads falsely identified as unique due to the few missing bases probably removed during trimming, were discarded.

### 5.5. Minor nucleotide variant calling analysis

Variant calling was conducted using an in-house R script similarly as previously described^10^. Samples with failed forward or reverse reactions were excluded from MNV analysis. Nucleotides with a mean Phred score ≤30, nucleotides observed ≤3 in each position, and nucleotide positions with <100x coverage were omitted from the analysis. All nucleotide variants for each position were counted from forward and reverse reactions separately. The most frequent variant in a position was designated Major while the second most frequent variant was designated MNV. If identified MNVs differed between forward and reverse reactions, the variant in the reaction with the highest coverage was used. A filtering step discarded MNVs with a frequency ≤1% within a sample. Detected MNVs of targeted HPV types found in the non-coding region (NCR) were manually investigated, discarding those present in homopolymeric regions (nucleotide repeated ≥ 5 times). Called MNVs were used in the mutational signature analysis classifying MNVs as either C>A, C>G, C>T, T>A, T>C, or T>G substitutions and further into 96 trinucleotide contexts and the proportional number of each substitution in each trinucleotide context per sample was calculated.

### 5.6. Reproducibility and repeatability analysis

The reproducibility and repeatability of the method were assessed using within-run and/or between-run replicates. After removing samples that failed to meet the filtering criteria, eight between-run and one within-run pairs underwent variant calling and integration analysis. Consensus sequences were generated by combining the forward and reverse reactions and calling the nucleotide with the highest coverage in each position (min. 20x). The consensus sequences and MNVs identified between technical replicate pairs were then compared against each other.

## Data Availability

The data presented in this article are not readily available because of the principles and conditions set out in the General Data Protection Regulation (GDPR), with additional national legal basis as per the Regulations on population-based health surveys and ethical approval from the Norwegian Regional Committee for Medical and Health Research Ethics (REC). Requests to access the data should be directed to the corresponding authors.

## 6. List of abbreviations

HPV: Human papillomavirus
SARS-CoV-2: Severe acute respiratory syndrome coronavirus 2
MNV: Minor nucleotide variant
TaMe-seq: Tagmentation-assisted multiplex PCR enrichment sequencing protocol
HR: High risk

## 7. Declarations

### 7.1. Ethics approval and consent to participate

The study involving HPV samples was approved by the Regional Committee for Medical and Health Research Ethics, Oslo, Norway (REK 2017/447) and by Akershus University Hospital’s Data Protection Official (2017-109). The study involving SARS-CoV-2 samples was approved by Akershus University Hospital’s Data Protection Official (20/06980) and the Regional Committee for Medical and Health Research Ethics, Oslo, Norway (REK 159268).

### 7.2. Consent for publication

Not applicable.

### 7.6. Authors’ contributions

AHL and MS designed and performed the experiments, analysed the results and drafted the manuscript text. JMC contributed to the experiments and analysis of the results. IKC managed the HPV sample material, contributed to the study design and result interpretation. TBR and OHA contributed to the study design, data analysis and result interpretation. HVA managed the SARS-CoV-2 sample material, contributed to the study design and result interpretation. All authors contributed to writing and approved the final version of the manuscript.

## 7.7. Acknowledgements

We would like to thank the International HPV Reference Centre, Karolinska Institute, Sweden, for sample material from the 2019 HPV proficiency panel used as controls in this study. We would also like to thank Pekka Ellonen and Harri Kangas at the Finish Institute of Molecular Medicine for invaluable help during the development of the updated TaME-seq2. The sequencing service was provided by the Norwegian Sequencing Centre (https://www.sequencing.uio.no), a national technology platform hosted by Oslo University Hospital and the University of Oslo supported by the Research Council of Norway and the South-Eastern Regional Health Authority.

## References

1. Bosch FX, Lorincz A, Muñoz N, Meijer Cjlm, Shah K v. The causal relation between human papillomavirus and cervical cancer. J Clin Pathol. 2002;55(4):244. doi:10.1136/JCP.55.4.244

2. Mirabello L, Yeager M, Yu K, et al. HPV16 E7 Genetic Conservation Is Critical to Carcinogenesis. Cell. 2017;170(6):1164-1174.e6. doi:10.1016/j.cell.2017.08.001

3. Zhu B, Xiao Y, Yeager M, et al. Mutations in the HPV16 genome induced by APOBEC3 are associated with viral clearance. Nature Communications 2020 11:1. 2020;11(1):1–12. doi:10.1038/s41467-020-14730-1

4. Aldunate F, Echeverría N, Chiodi D, et al. Resistance-associated substitutions and response to treatment in a chronic hepatitis C virus infected-patient: an unusual virological response case report. BMC Infect Dis. 2021;21(1):1–6. doi:10.1186/S12879-021-06080-0/FIGURES/2

5. Kyeyune F, Gibson RM, Nanky I, et al. Low-Frequency Drug Resistance in HIV-Infected Ugandans on Antiretroviral Treatment Is Associated with Regimen Failure. Antimicrob Agents Chemother. 2016;60(6):3380–3397. doi:10.1128/AAC.00038-16

6. Armero A, Berthet N, Avarre JC. Intra-Host Diversity of SARS-Cov-2 Should Not Be Neglected: Case of the State of Victoria, Australia. Viruses. 2021;13(1). doi:10.3390/V13010133

7. Tonkin-Hill G, Martincorena I, Amato R, et al. Patterns of within-host genetic diversity in SARS-COV-2. Elife. 2021;10. doi:10.7554/ELIFE.66857

8. Burk RD, Chen Z, Saller C, et al. Integrated genomic and molecular characterization of cervical cancer. Nature 2017 543:7645. 2017;543(7645):378–384. doi:10.1038/nature21386

9. McBride AA, Warburton A. The role of integration in oncogenic progression of HPV-associated cancers. PLoS Pathog. 2017;13(4):e1006211. doi:10.1371/JOURNAL.PPAT.1006211

10. Lagström S, Umu SU, Lepistö M, et al. TaME-seq: An efficient sequencing approach for characterisation of HPV genomic variability and chromosomal integration. Scientific Reports 2019 9:1. 2019;9(1):1–12. doi:10.1038/s41598-018-36669-6

11. Cullen M, Boland JF, Schiffman M, et al. Deep sequencing of HPV16 genomes: A new high-throughput tool for exploring the carcinogenicity and natural history of HPV16 infection. Papillomavirus Res. 2015;1:3–11. doi:10.1016/J.PVR.2015.05.004

12. Holmes A, Lameiras S, Jeannot E, et al. Mechanistic signatures of HPV insertions in cervical carcinomas. NPJ Genom Med. 2016;1:16004. doi:10.1038/NPJGENMED.2016.4

13. Escobar-Escamilla N, Ramírez-González JE, Castro-Escarpulli G, Díaz-Quiñonez JA. Utility of high-throughput DNA sequencing in the study of the human papillomaviruses. Virus Genes. 2018;54(1):17–24. doi:10.1007/S11262-017-1530-3

14. Lagström S, Løvestad AH, Umu SU, et al. HPV16 and HPV18 type-specific APOBEC3 and integration profiles in different diagnostic categories of cervical samples. Tumour Virus Res. 2021;12:200221. doi:10.1016/J.TVR.2021.200221

15. Lagström S, van der Weele P, Rounge TB, Christiansen IK, King AJ, Ambur OH. HPV16 whole genome minority variants in persistent infections from young Dutch women. J Clin Virol. 2019;119:24–30. doi:10.1016/J.JCV.2019.08.003

16. Tropé A, Sjøborg KD, Nygård M, et al. Cytology and Human Papillomavirus Testing 6 to 12 Months after ASCUS or LSIL Cytology in Organized Screening To Predict High-Grade Cervical Neoplasia between Screening Rounds. J Clin Microbiol. 2012;50(6):1927. doi:10.1128/JCM.00265-12

17. Tropé A, Sjøborg K, Eskild A, et al. Performance of human papillomavirus DNA and mRNA testing strategies for women with and without cervical neoplasia. J Clin Microbiol. 2009;47(8):2458–2464. doi:10.1128/JCM.01863-08

18. Eklund C, Mühr LSA, Lagheden C, Forslund O, Robertsson KD, Dillner J. The 2019 HPV Labnet international proficiency study: Need of global Human Papillomavirus Proficiency Testing. Journal of Clinical Virology. 2021;141:104902. doi:10.1016/J.JCV.2021.104902

19. Løvestad AH, Jørgensen SB, Handal N, Ambur OH, Aamot H v. Investigation of intra-hospital SARS-CoV-2 transmission using nanopore whole-genome sequencing. Journal of Hospital Infection. 2021;111:107–116. doi:10.1016/j.jhin.2021.02.022

20. artic-network/artic-ncov2019: ARTIC nanopore protocol for nCoV2019 novel coronavirus. Accessed October 28, 2022. https://github.com/artic-network/artic-ncov2019

21. Kozich JJ, Westcott SL, Baxter NT, Highlander SK, Schloss PD. Development of a dual-index sequencing strategy and curation pipeline for analyzing amplicon sequence data on the MiSeq Illumina sequencing platform. Appl Environ Microbiol. 2013;79(17):5112–5120. doi:10.1128/AEM.01043-13

22. Kim D, Langmead B, Salzberg SL. HISAT: a fast spliced aligner with low memory requirements. Nature Methods 2015 12:4. 2015;12(4):357–360. doi:10.1038/nmeth.3317

23. van Doorslaer K, Tan Q, Xirasagar S, et al. The Papillomavirus Episteme: a central resource for papillomavirus sequence data and analysis. Nucleic Acids Res. 2013;41(Database issue):D571. doi:10.1093/NAR/GKS984

24. Roberts SA, Lawrence MS, Klimczak LJ, et al. An APOBEC cytidine deaminase mutagenesis pattern is widespread in human cancers. Nature Genetics 2013 45:9. 2013;45(9):970–976. doi:10.1038/ng.2702

25. Korbie DJ, Mattick JS. Touchdown PCR for increased specificity and sensitivity in PCR amplification. Nat Protoc. 2008;3(9):1452–1456. doi:10.1038/NPROT.2008.133

26. Smits N, Rasmussen J, Bodea GO, et al. No evidence of human genome integration of SARS-CoV-2 found by long-read DNA sequencing. Cell Rep. 2021;36(7):109530. doi:10.1016/J.CELREP.2021.109530

27. Løvestad AH, Repesa A, Costanzi JM, et al. Differences in integration frequencies and APOBEC3 profiles of five high-risk HPV types adheres to phylogeny. Tumour Virus Res. 2022;14:200247. doi:10.1016/J.TVR.2022.200247

28. Oliveira CM de, Bravo IG, Souza NCS e., et al. High-level of viral genomic diversity in cervical cancers: A Brazilian study on human papillomavirus type 16. Infection, Genetics and Evolution. 2015;34:44–51. doi:10.1016/J.MEEGID.2015.07.002

29. Hirose Y, Onuki M, Tenjimbayashi Y, et al. Within-Host Variations of Human Papillomavirus Reveal APOBEC Signature Mutagenesis in the Viral Genome. J Virol. 2018;92(12):17–18. doi:10.1128/JVI.00017-18

30. Arroyo-Mühr LS, Lagheden C, Hultin E, et al. The HPV16 Genome Is Stable in Women Who Progress to In Situ or Invasive Cervical Cancer: A Prospective Population-Based Study. Cancer Res. 2019;79(17):4532–4538. doi:10.1158/0008-5472.CAN-18-3933

31. Pachetti M, Marini B, Benedetti F, et al. Emerging SARS-CoV-2 mutation hot spots include a novel RNA-dependent-RNA polymerase variant. J Transl Med. 2020;18(1):1–9. doi:10.1186/S12967-020-02344-6/FIGURES/4

32. Kebschull JM, Zador AM. Sources of PCR-induced distortions in high-throughput sequencing data sets. Nucleic Acids Res. 2015;43(21):e143. doi:10.1093/NAR/GKV717

