## Supplementary file for "TaME-seq2: Tagmentation-assisted multiplex PCR enrichment sequencing for viral genomic profiling"

Table S1. Overview of cytology-based and/or histology classification of clinical samples positive for HPV51, 52, and 59. (LSIL – low-grade intraepithelial lesion, ASC-US - atypical squamous cells of undetermined significance, HSIL- high-grade intraepithelial lesion, ASC-H - Atypical squamous cells- cannot exclude high-grade squamous intraepithelial lesion, CIN1- Cervical intraepithelial neoplasia grade 1, CIN2-Cervical intraepithelial neoplasia grade 2, ND-not determined, NR-not relevant)

| **Sample name** | **Cytology-based classification** | **Histology-based classification** |
| --- | --- | --- |
| **HPV51** | | |
| 1-HPV51 | HSIL | ND |
| 2-HPV51 | ASC-US | ND |
| 3-HPV51 | LSIL | ND |
| 4-HPV51 | NR | CIN2 |
| 5-HPV51 | Normal morphology | ND |
| 6-HPV51 | ASC-US | ND |
| 7-HPV51 | HSIL | ND |
| 8-HPV51 | NR | CIN2 |
| 9-HPV51 | Normal benign cells | ND |
| 10-HPV51 | HSIL | ND |
| 11-HPV51 | Normal morphology | ND |
| 12-HPV51 | Severe dysplasia, carcinoma *in situ* in squamous epithelium cells | ND |
| **HPV52** | | |
| 1-HPV52 | NR | CIN2 |
| 2-HPV52 | HSIL | ND |
| 3-HPV52 | ASC-US | ND |
| 4-HPV52 | ASC-US | ND |
| 5-HPV52 | Normal morphology | ND |
| 6-HPV52 | Normal morphology | ND |
| 7-HPV52 | Normal morphology | ND |
| 8-HPV52 | HSIL | ND |
| 9-HPV52 | Normal morphology | ND |
| 10-HPV52 | Irregular simple columnar epithelium, malignancy uncertain | ND |
| 11-HPV52 | Severe dysplasia, carcinoma *in situ* in squamous epithelium cells | ND |
| 12-HPV52 | NA | CIN2 |
| **HPV59** | | |
| 1-HPV59 | HSIL | ND |
| 2-HPV59 | HSIL | ND |
| 3-HPV59 | Normal morphology | ND |
| 4-HPV59 | LSIL | ND |
| 5-HPV59 | Normal morphology | ND |
| 6-HPV59 | HSIL | ND |
| 7-HPV59 | LSIL | ND |
| 8-HPV59 | ASC-US | ND |
| 9-HPV59 | NR | CIN2 |
| 10a-HPV59 | Severe dysplasia, carcinoma *in situ* in squamous epithelium cells | ND |
| 10b-HPV59 | Severe dysplasia, carcinoma *in situ* in squamous epithelium cells | ND |
| 11-HPV59 | Normal morphology | ND |

Table S2. Read counts and sequencing statistics of HPV positive cervical cell samples, HPV harboring plasmids (Equalis proficiency panel) and SARS-CoV-2 positive clinical samples (NA-not applicable).

| **Sample** | **Sample type** | **Raw reads** | **Trimmed reads** | **Reads mapped to target HPV** | **% Reads mapped**  **to target HPV** | **Mean coverage** | **% of genome covered by minimum** | | **Nº of variable sites in samples passing**  **300x mean coverage** |
| --- | --- | --- | --- | --- | --- | --- | --- | --- | --- |
|  |  |  |  |  |  |  | **10x** | **100x** |  |
| **HPV51** | | | | | | | | | |
| 8-HPV51 | LBC | 37711658 | 12600990 | 8368861 | 22.19% | 71702.37 | 99.95% | 99.83% | 7 |
| 1-HPV51 | LBC | 27052536 | 7787978 | 6790878 | 25.10% | 62943.62 | 99.95% | 99.83% | 10 |
| 12-HPV51 | LBC | 22996764 | 9956824 | 5355924 | 23.29% | 52417.84 | 99.83% | 99.78% | 4 |
| 5-HPV51 | LBC | 13598120 | 4489808 | 3825525 | 28.13% | 36921.88 | 98.63% | 86.92% | NA |
| 4-HPV51 | LBC | 21891556 | 10772478 | 2767160 | 12.64% | 27662.42 | 99.72% | 99.56% | 13 |
| 3-HPV51 | LBC | 4152018 | 2556750 | 2678514 | 64.51% | 27339.49 | 99.74% | 97.11% | NA |
| 6-HPV51 | LBC | 14391794 | 5171188 | 642780 | 4.47% | 6176.23 | 99.82% | 96.88% | 6 |
| 9-HPV51 | LBC | 30044338 | 12316120 | 611487 | 2.04% | 6078.92 | 99.80% | 97.52% | 53 |
| 11-HPV51 | LBC | 22848534 | 9951756 | 470212 | 2.06% | 4655.98 | 99.78% | 91.48% | 24 |
| 2-HPV51 | LBC | 31596160 | 13368302 | 257352 | 0.81% | 2695.61 | 99.80% | 91.18% | 45 |
| 10-HPV51 | LBC | 29971500 | 11437710 | 34241 | 0.11% | 333.87 | 75.40% | 40.91% | 29 |
| 7-HPV51 | LBC | 16462424 | 7190730 | 8 | 0.00% | 0.08 | 0.00% | 0.00% | NA |
| pEq25-HPV51 | Plasmid. single pHPV51 | 11431124 | 4636648 | 62263 | 0.54% | 615.63 | 97.53% | 57.59% | NA |
| pEq14-HPV51 | Plasmid. mixture of  pHPV6/16/18/51 | 9488828 | 4024620 | 10076 | 0.11% | 98.27 | 74.96% | 19.75% | NA |
| H20-HPV51 | Water | 59776 | 45372 | 0 | 0.00% | NA | NA | NA | NA |
| **HPV52** | | | | | | | | | |
| 1-HPV52 | LBC | 24714832 | 9501798 | 8784246 | 35.54% | 82704.09 | 100.00% | 100.00% | 3 |
| 10-HPV52 | LBC | 12443322 | 6291884 | 4690774 | 37.70% | 50735.10 | 100.00% | 100.00% | 3 |
| 12-HPV52 | LBC | 10752286 | 5952656 | 801631 | 7.46% | 8500.72 | 100.00% | 100.00% | 12 |
| 11-HPV52 | LBC | 10067914 | 4934694 | 583587 | 5.80% | 5834.53 | 100.00% | 99.95% | 25 |
| 7-HPV52 | LBC | 10679198 | 5099056 | 517462 | 4.85% | 5444.72 | 99.99% | 99.71% | 22 |
| 5-HPV52 | LBC | 7696962 | 3960162 | 163600 | 2.13% | 1720.31 | 100.00% | 93.15% | 38 |
| 9-HPV52 | LBC | 10000182 | 6311054 | 57901 | 0.58% | 606.93 | 87.98% | 64.63% | 31 |
| 8-HPV52 | LBC | 308764 | 275034 | 24220 | 7.84% | 287.86 | 76.87% | 52.58% | NA |
| 6-HPV52 | LBC | 10338864 | 6334572 | 12442 | 0.12% | 131.45 | 71.30% | 36.54% | NA |
| 2-HPV52 | LBC | 10449560 | 6377802 | 4939 | 0.05% | 49.17 | 37.55% | 11.17% | NA |
| 3-HPV52 | LBC | 5832794 | 4133402 | 2108 | 0.04% | 23.06 | 31.43% | 7.18% | NA |
| 4-HPV52 | LBC | 12070702 | 5747126 | 208 | 0.00% | 1.51 | 5.46% | 0.00% | NA |
| pEq21-HPV52 | Plasmid. single pHPV52 | 6166886 | 4335382 | 21191 | 0.34% | 243.65 | 73.19% | 43.54% | NA |
| pEq32-HPV52 | Plasmid mixture of  pHPV39/45/52/68b | 5246422 | 3609510 | 183904 | 3.51% | 1906.88 | 96.13% | 80.31% | NA |
| H20-HPV52 | Water | 79988 | 68264 | 0 | 0.00% | NA | 100.00% | 100.00% | NA |
| **HPV59** | | | | | | | | | |
| 7-HPV59 | LBC | 19422168 | 7178738 | 7154100 | 36.83% | 61721.02 | 100.00% | 100.00% | 9 |
| 4-HPV59 | LBC | 22772450 | 7510366 | 7676229 | 33.71% | 51920.88 | 100.00% | 100.00% | 5 |
| 8-HPV59 | LBC | 31342168 | 7127906 | 6771112 | 21.60% | 35452.37 | 100.00% | 100.00% | 30 |
| 6-HPV59 | LBC | 3959998 | 2181194 | 1537875 | 38.84% | 16585.57 | 100.00% | 97.85% | 12 |
| 10b-HPV59 | LBC | 3974280 | 2581018 | 840639 | 21.15% | 9436.97 | 91.29% | 89.97% | 27 |
| 10a-HPV59 | LBC | 1806050 | 1114120 | 264352 | 14.64% | 2976.90 | 91.22% | 88.48% | 32 |
| 9-HPV59 | LBC | 11323896 | 7423128 | 34478 | 0.30% | 364.16 | 76.30% | 46.63% | 26 |
| 2-HPV59 | LBC | 1663864 | 1099422 | 14289 | 0.86% | 152.70 | 82.62% | 41.67% | NA |
| 5-HPV59 | LBC | 20000 | 14522 | 5231 | 26.16% | 61.57 | 87.45% | 21.49% | NA |
| 1-HPV59 | LBC | 518056 | 391140 | 2597 | 0.50% | 31.28 | 46.35% | 6.65% | NA |
| 3-HPV59 | LBC | 6821068 | 4083508 | 1047 | 0.02% | 12.35 | 20.48% | 2.10% | NA |
| 11-HPV59 | LBC | 6108 | 4794 | 5 | 0.08% | 0.06 | 0.00% | 0.00% | NA |
| pEq26-HPV59 | Plasmid. single pHPV59 | 9237822 | 5844454 | 24631 | 0.27% | 253.76 | 71.87% | 40.16% | NA |
| pEq23-HPV59 | Plasmid mixture of pHPV35/56/59/68a | 9726618 | 6484628 | 541287 | 5.57% | 6088.65 | 100.00% | 98.52% | NA |
| H20-HPV59 | Water | 183412 | 157842 | 0 | 0.00% | NA | NA | NA | NA |
| **SARS-CoV-2** | | | | | | | | | |
| **Sample** | **Sample type** | **Raw reads** | **Trimmed reads** | **Reads mapped to target HPV** | **% Reads mapped**  **to target HPV** | **Mean coverage** | **% of genome covered by minimum** | | **Nº of variable sites in samples passing**  **300x mean coverage** |
|  |  |  |  |  |  |  | **10x** | **100x** |  |
| 3_3-CoV | cDNA | 49700332 | 20016968 | 11918481 | 23.98 % | 31475.46 | 82.56 % | 65.07 % | 24 |
| 1_6-CoV | cDNA | 19011470 | 8751918 | 6047458 | 31.81 % | 17826.32 | 97.80 % | 91.65 % | 73 |
| 3_2-CoV | cDNA | 14013914 | 4288876 | 3627061 | 25.88 % | 10347.62 | 89.51 % | 82.94 % | 177 |
| 2_6-CoV | cDNA | 23452764 | 13179208 | 2812113 | 11.99 % | 7660.54 | 67.96 % | 57.32 % | 78 |
| 4_1-CoV | cDNA | 13621222 | 7492924 | 1812212 | 13.30 % | 4821.64 | 88.01 % | 80.07 % | 215 |
| 3_4-CoV | cDNA | 16118934 | 10843988 | 1369827 | 8.50 % | 3703.35 | 67.66 % | 55.29 % | 98 |
| 3_7-CoV | cDNA | 14766420 | 10156544 | 824458 | 5.58 % | 2397.13 | 85.15 % | 64.05 % | 148 |
| 3_5-CoV | cDNA | 3894808 | 2597890 | 144083 | 3.70 % | 431.09 | 60.36 % | 42.23 % | NA |
| 2_5-CoV | cDNA | 15277072 | 14425932 | 55120 | 0.36 % | 162.86 | 57.86 % | 29.71 % | NA |
| 3_8-CoV | cDNA | 6626066 | 4174742 | 61254 | 0.92 % | 152.44 | 42.37 % | 21.48 % | NA |
| 2_4-CoV | cDNA | 17666524 | 16158748 | 45700 | 0.26 % | 139.40 | 37.99 % | 15.52 % | NA |
| 2_8-CoV | cDNA | 86835886 | 75779800 | 30672 | 0.04 % | 82.03 | 27.58 % | 6.93 % | NA |
| 2_1-CoV | cDNA | 24489306 | 23169198 | 23979 | 0.10 % | 64.02 | 35.25 % | 11.83 % | NA |
| 2_2-CoV | cDNA | 21484810 | 20002254 | 17771 | 0.08 % | 51.15 | 36.43 % | 11.26 % | NA |
| 1_5-CoV | cDNA | 17476402 | 16063872 | 21568 | 0.12 % | 49.73 | 21.23 % | 8.18 % | NA |
| 1_3-CoV | cDNA | 19152872 | 18421058 | 11551 | 0.06 % | 34.39 | 40.42 % | 8.85 % | NA |
| 1_2-CoV | cDNA | 17358898 | 15307590 | 8040 | 0.05 % | 23.41 | 30.37 % | 5.52 % | NA |
| 2_7-CoV | cDNA | 72299428 | 65464234 | 7233 | 0.01 % | 16.68 | 15.59 % | 2.07 % | NA |
| 1_8-CoV | cDNA | 33776278 | 31288986 | 729 | 0.00 % | 1.85 | 5.13 % | 0.21 % | NA |
| 1_1-CoV | cDNA | 24009794 | 23263926 | 652 | 0.00 % | 1.57 | 1.90 % | 0.24 % | NA |
| 1_4-CoV | cDNA | 1114888 | 994682 | 463 | 0.04 % | 1.13 | 3.17 % | 0.00 % | NA |
| 1_7-CoV | cDNA | 16238506 | 14907718 | 227 | 0.00 % | 0.56 | 0.26 % | 0.25 % | NA |
| 3_1-CoV | cDNA | 1083412 | 986258 | 92 | 0.01 % | 0.33 | 0.44 % | 0.00 % | NA |

Table S3. Location of the integration breakpoints found in duplicates of 10-HPV59 sample and number of unique discordant read pairs and junction reads at the integration breakpoints.

| **Sample** | **Reaction** | **HPV** | | **Human breakpoint**  **(Grch38/hg38)** | **Unique discordant read pairs** | **Unique junction reads** |
| --- | --- | --- | --- | --- | --- | --- |
|  |  | **Breakpoint** | **ORF** |  |  |  |
| 10a-HPV59 | F | 3149 | E2 | Chr2:225998434 | 6 | 0 |
| 10b-HPV59 | F | 3149 | E2 | Chr2:225998437 | 8 | 0 |
| 10b-HPV59 | F | 3148 | E2 | Chr2:225998384 | 0 | 12 |
| 10a-HPV59 | R | 4031 | E5 | Chr2226171560 | 0 | 22 |
| 10b-HPV59 | R | 4031 | E5 | Chr2226171552 | 0 | 3 |
| 10b-HPV59 | R | 4031 | E5 | Chr2226171558 | 0 | 3 |
| 10a-HPV59 | R | 4031 | E5 | Chr2226171560 | 0 | 43 |
| 10b-HPV59 | R | 4037 | E5 | Chr2226171560 | 0 | 5 |

Table S4. Number of minor nucleotide variants (MNVs) grouped by HPV gene, identified in run 1 and run 2.

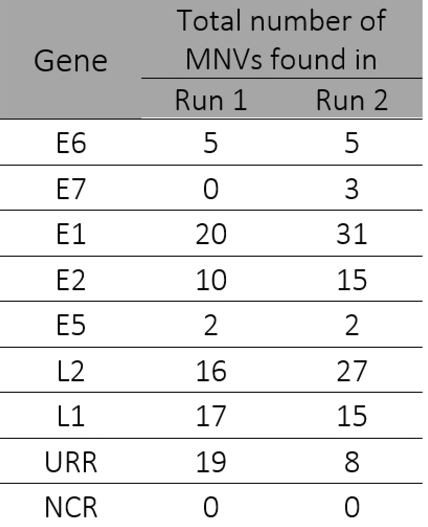

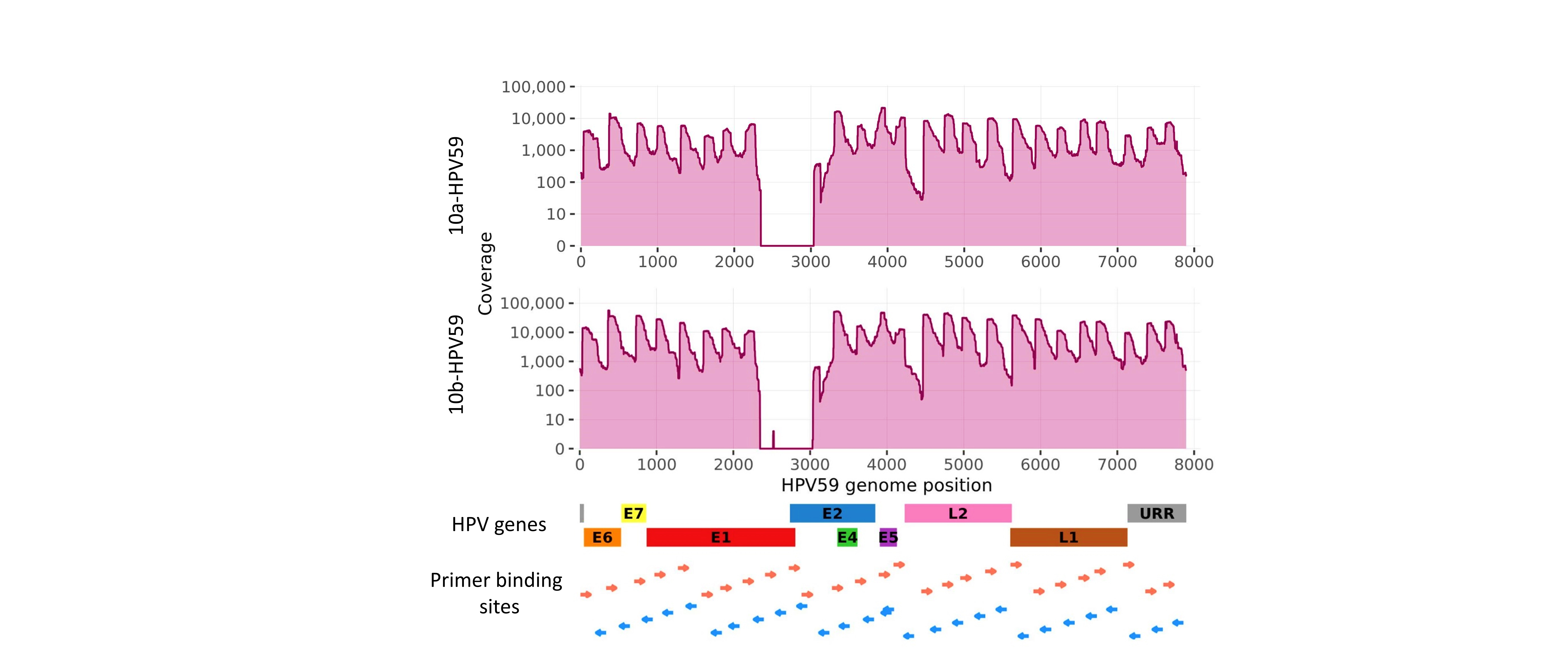

Figure S1. HPV genome sequencing coverage in replicates 10a-HPV59 and 10b-HPV59.

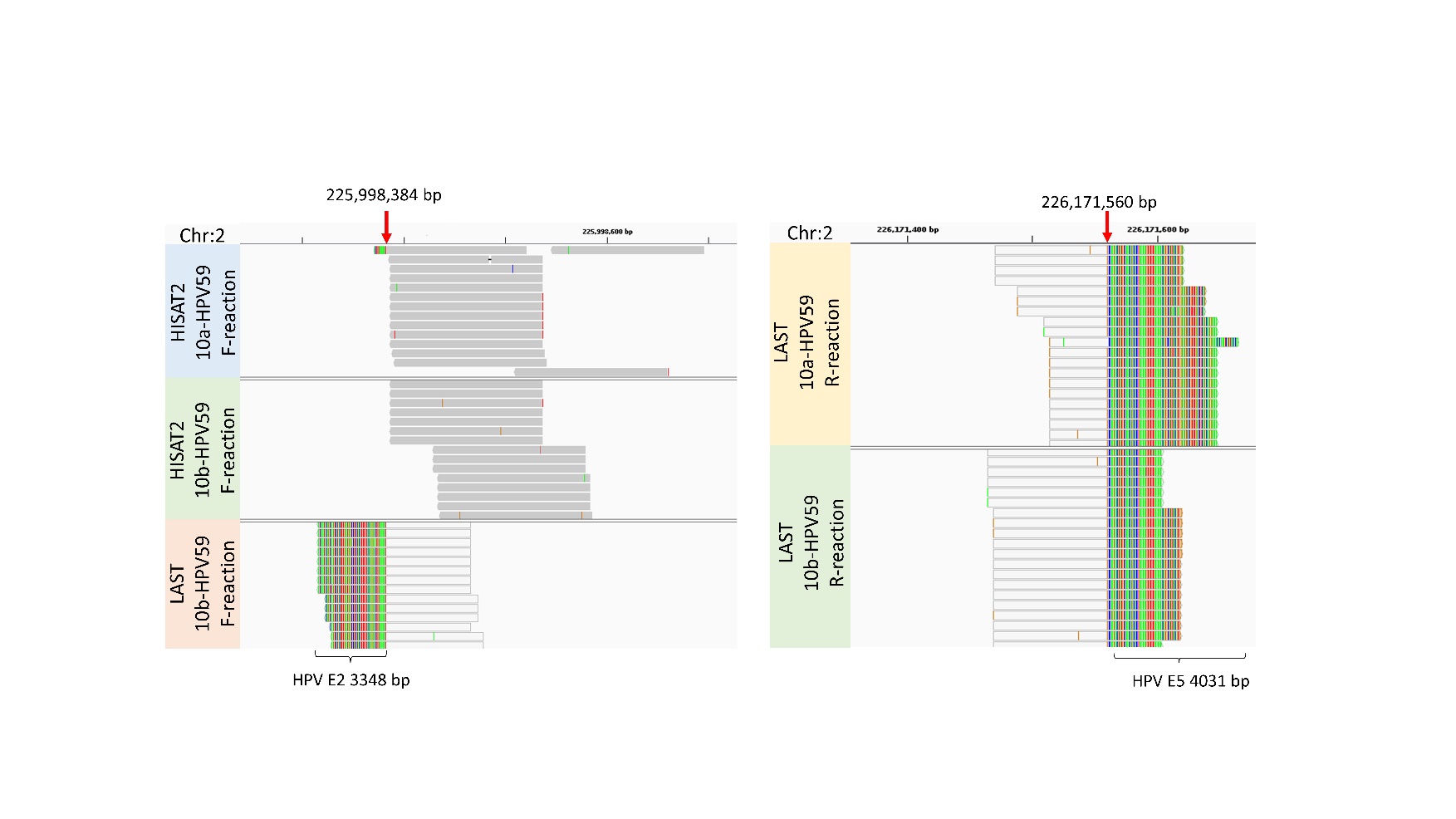

Figure S2. IGV visualization of HISAT2 and LAST alignments of 10a-HPV59 and 10b-HPV59 to human genome indicating HPV-human integration breakpoints. (a) Integration breakpoint between chromosome 2 (GRCh38/hg38) and HPV59 E2. (b) Integration breakpoint between chromosome 2 and HPV59 E5. HISAT2 reads are presented with dark gray color. Parts of the LAST reads mapping to human are light gray, while parts mapping to HPV59 are multi-colored. Red arrows point to the exact breakpoint positions.

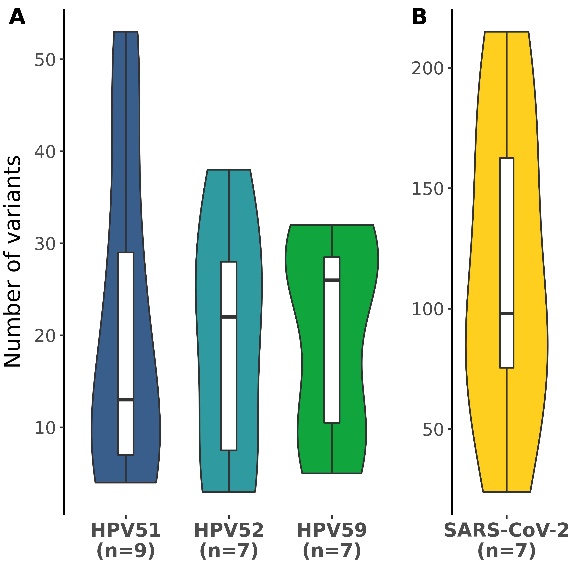

Figure S3. Number of minor nucleotide variants presented as violin plots across the different virus types, **A)** HPV52, HPV52, and HPV59; **B)** SARS-CoV-2. Violin plot shows the probable density of the data, using kernel density estimation. Box-and-whisker plots are added to show the median number (horizontal line), 25% and 75% percentiles (box), minimum and maximum values (whiskers).

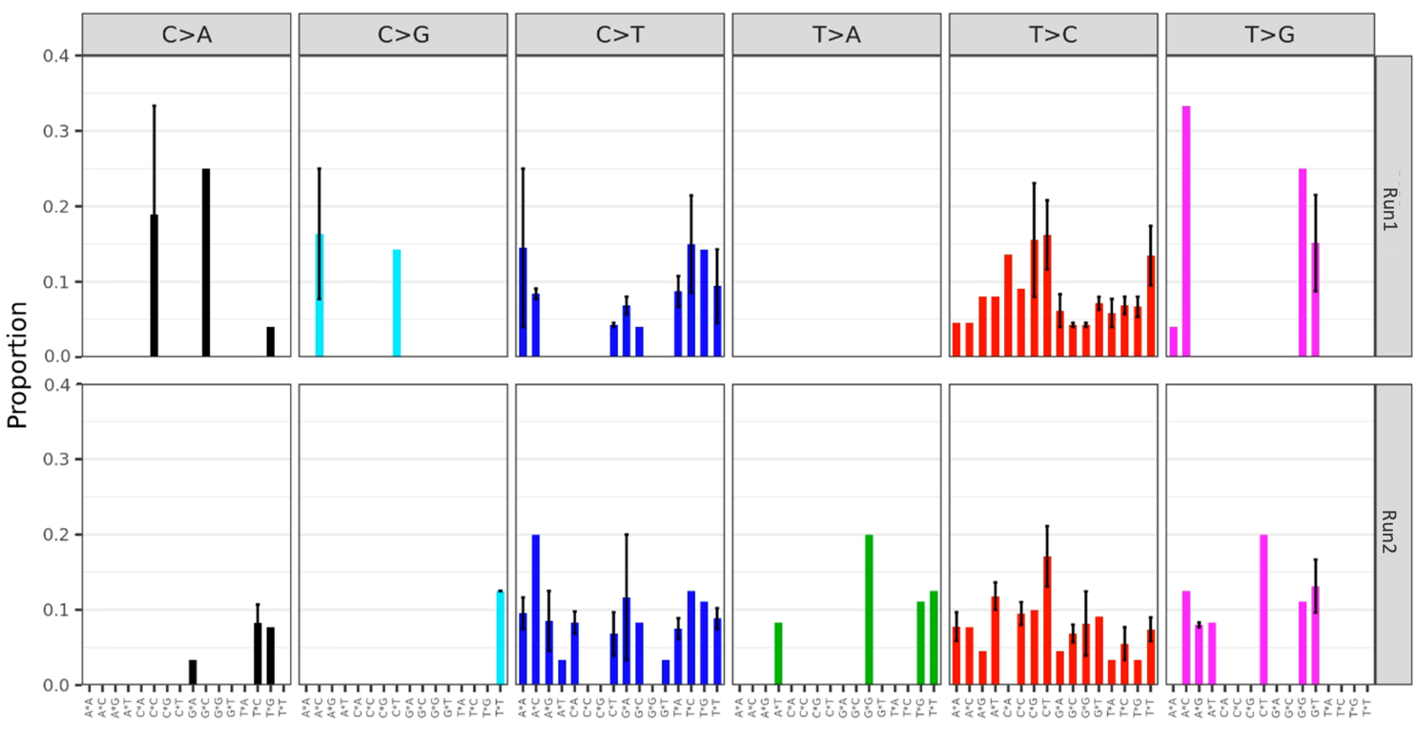

Figure S4. Mutational signatures found between technical replicates in the two sequencing runs. The mean proportion of 96 trinucleotide substitution types is shown below the plots across the different diagnostic categories. Error bars represent the standard error of the mean.
